# Effects of climatic variables on dengue incidence in Cali

**DOI:** 10.1101/2024.05.01.24306676

**Authors:** Mauricio Frieri, Marisol Gordillo, Lilian S. Sepúlveda

**Affiliations:** Faculty of Engineering and Basic Sciences, Universidad Autónoma de Occidente, Cali, Colombia

**Keywords:** Dengue, Negative binomial regression model, random forest regression, climatic variables

## Abstract

In this work we studied the relationship between dengue incidence in Cali and the climatic variables that are known to have an impact on the mosquito and were available (precipitation, relative humidity, minimum, mean, and maximum temperature). Since the natural processes of the mosquito imply that any changes on climatic variables need some time to be visible on the dengue incidence, a lagged correlation analysis was done in order to choose the predictor variables of count regression models. A Principal Component Analysis was done to reduce dimensionality and study the correlation among the climatic variables. Finally, aiming to predict the monthly dengue incidence, three different regression models were constructed and compared using de Akaike information criterion. The best model was the negative binomial regression model, and the predictor variables were mean temperature with a 3-month lag and mean temperature with a 5-month lag as well as their interaction. The other variables were not significant on the models. And interesting conclusion was that according to the coefficients of the regression model, a 1°C increase in the monthly mean temperature will reflect as a 45% increase in dengue incidence after 3 months. The rises to a 64% increase after 5 months.

**Author Summary:** Dengue is transmitted by the bite of an infected mosquito, and mosquitoes, in turn, are affected by climatic conditions. In this work studied the relationship between dengue incidence in Cali and climatic variables, namely precipitation, relative humidity, minimum temperature, mean temperature, and maximum temperature using statistical methods. Since this is a natural and biological process, the changes in climatic conditions need time to have a visible effect on dengue incidence, hence we identified the significant climatic variables and the time they take to have a visible effect on dengue incidence. Then, we created three different models for predicting dengue incidences using the lagged variables and picked the best one. We concluded that the most critical variable is mean temperature with a 3- and 5-month lag. We also found that a 1°C increase in the monthly mean temperature will reflect as a 45% increase in dengue incidence after 3 months. The rises to a 64% increase after 5 months.

## 1. Introduction

Dengue is the viral disease transmitted by *Aedes aegypti* with the biggest worldwide spreading. It’s common in warm and humid areas all around the world. It’s transmitted by the biting of infected mosquitoes, and it can cause high fever, nausea, vomiting, rash and pain in different parts of the body (eyes, muscles, joints or bones). When severe, it may also cause internal bleeding and even death [1].

Every year around 50 million dengue cases are registered, and approximately 2.500 million people live in countries where this disease is endemic [2]. Furthermore, dengue affects Cali, a city in the southwest of Colombia, chronically. In fact, yearly mean rates of dengue in Cali reach almost 100 cases per 100.000 inhabitants [3].

On the other hand, it’s well known that climatic conditions play a big part on the reproduction and maturation of *Aedes aegipti.* Warm temperature shortens the incubation time of the larvae, while precipitation directly affects the availability of breeding places [4–6] Also, temperature exerts a significant influence and regulates the development of mosquitoes [7]. However, excesses of precipitation might imply a reduction of the population since nor larvae nor pupae survive in these conditions [8–11]. Additionally, El Niño phenomenon (ELSO) is related with dengue incidence and the effects take several months to be reflected on it depending on the region [12–14].

Concerning the modeling of dengue, various count regression models have been successfully used [15–18]. Amongst them there is the negative binomial model since it, unlike the Poisson model, doesn’t assume equality between the mean and the variance. Using this type of models [19–22] found that temperature and relative humidity are important variables for predicting dengue incidence.

In Cali, diverse intervention measures have been implemented. In 2022, the activities carried out include drain inspections, household visits for eliminating hatcheries, control campaigns, biological control, and fumigation. For 2023, the Public Health Department of Cali will begin its work with initiatives aiming at education and prevention [23]. Nevertheless, it is still crucial to continue the research on the impact of climatic variables on dengue incidence to prevent endemic spikes and adjust control strategies.

Because of the way dengue is propagated, focusing control strategies on the vector is essential, which generally imply intensive and expensive efforts. Hence, this control schemes must be implemented strategic fashion in order to maximize their efficiency and minimize costs. Because of this, the study of dengue incidence in relation with climatic variables associated with the reproduction and development of *Aedes aegypti* should not be disregarded.

The main goal of this research consisted of analyzing the relationship and delay between climatic variables and dengue incidence. We sought to describe and quantify how the climatic patterns are directly linked with dengue incidence throughout statistical analysis. Data of Cali was selected because this city presents one of the highest dengue infection rates in Colombia.

## 2. Materials and methods

### 2.1. Area of study

Cali, the third biggest city in Colombia, is the capital of the department of Valle del Cauca. It’s located on the south of the department, on the valley formed by the western and central branches of the Andes Mountain range, at a mean altitude of 1000 meters above sea level. Cali is also located on the valley of Cauca River, the second most important of Colombia. Cali’s geographical coordinates are 3°27’00’’N 76°32’00’’O. Cali’s weather is warm and humid, with an average temperature of 24°C.

The weather in Cali es characterized for being warm and dry, classifying as tropical with dry summers according to the Köppen Climate Classification. The western branch of the mountain range plays an important role stopping the humid air currents coming from the Pacific Ocean, although the maritime breezes do indeed reach the city. The mountain range has an average altitude of 2000 meters above sea level at the north of the city, reaching up to 4100 meters on the south, where the highest pluviosity occurs. On average, annual precipitation lies from 900 mm on the driest areas up to 1800 on the rainiest, being 1483 mm the average over the majority of Cali’s metropolitan area. Mean temperature es 24.0°C (73.6°F). From December to February and July to Augusts it’s considered the dry season, while from March to May and September to November is the rainy season [24].

### 2.2. Data collection

For this study, data of monthly dengue cases provided by the Public Health Department of Cali was used, encompassing a time period from January of 2015 to December 2021. Similarly, from the Hydrology, Meteorology and Environmental Studies Institute (IDEAM) monthly data were collected for the following climatic variables: maximum, minimum and average temperature (°C), precipitation (mm) and relative humidity (%) on the same time period.

### 2.3. Statistical methods

#### 2.3.1. Random Forest Regression

The random forest regression technique was used for missing data imputation with python’s *sklearn* library on Google Collab. This machine learning method stands out for its efficiency on both classification and regression problems, although in this work we only care for regression problems. This method is based on the construction of a set of decision trees that will be used to predict continuous values. This technique is increasingly been used in diverse areas for the processing of big volumes of data [25,26].

As shown on Fig 1, the main idea of random forest regression is to create multiple different regression trees trained with different samples from the training set. Thus, each tree will make different individual predictions that then will be averaged to produce a single final prediction. This method tends to be better than using a single regression tree since the latter is highly sensible to the training data and so, the averaged prediction is more robust.

**Fig. 1.**
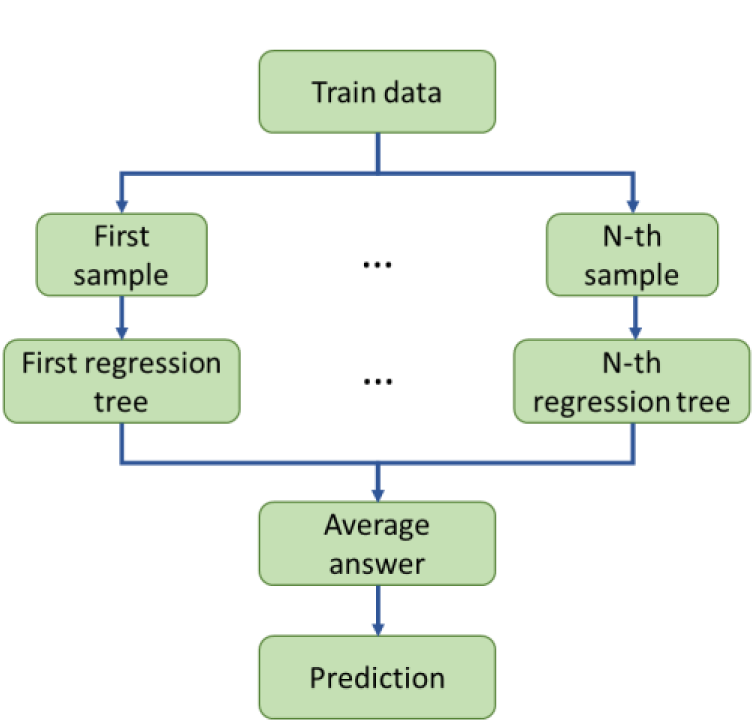
Random Forrest regression algorithm

Similarly, regression trees are a machine learning technique used for prediction problems. In other words, we want to predict the value of a continuous variable *y* using a set of predictor variables or covariables *X.* Hence, a tree is constructed such that on each node examples are split in a way were on each new node the variance of *y* is the least possible. This implies picking on every node a suitable covariable and splitting point *c.* This process is repeated until a stop criterion is reached which may depend on the depth of the tree or the number of examples on the nodes.

It’s essential to remark that each of the regression trees are different, and therefore return different predictions, because they are constructed from different random samples from the data base. This is called bagging and is used when working with noise data sets.

#### 2.3.2. Correlation analysis

Since the effect of climatic variables over the dengue incidence is not immediate, a lagged correlation analysis was made, a common tool of time series analysis, in order to select the variables and corresponding lag to be the predictors on the regression model. Subsequently, the number of dimensions was reduced by doing a principal component analysis (ACP) also avoiding multicollinearity issues.

#### 2.3.3. Principal component analysis

ACP is a useful statistical technique for synthesizing information or reducing the number of variables on a data set without losing important information. During ACP new variables are created called principal components, which are linear combinations of the original variables and, additionally, they are pairwise mutually independent.

In this work, ACP was used for two purposes. Firstly, as a data exploration tool in order to study the correlation between the climatic variables and, secondly, to reduce the amount of predictor variables in the statistical model. The similarities between these variables were studied and only one was chosen amongst the groups of homogeneous variables so it acts a representative, since multiple variables with like behavior is not needed. This analysis was made using SPSS.

#### 2.3.4. Regression models

Keeping in mind that dengue incidence is a count variable, three different regression models were considered for this project, namely, Poisson, negative binomial, and Poisson-inverse Gaussian. The Poisson model is one of the more classical ones for count data. It assumes that the data follow a Poisson distribution with the following probability distribution function:

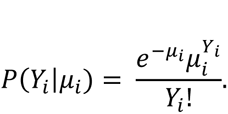

Here, *Y*_*i*_ is a random variable with mean and variance *µ*_*i*_. For constructing regression models, *µ*_*i*_ is expressed as a function of the independent variables throughout the natural log as a link function:

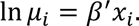

Similarly, the dependent variable is link, throughout the natural log, whit a linear function of the independent variables:

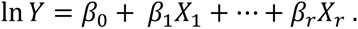

The negative binomial model is similar to the Poisson model with the key difference that it relaxes the assumption of equality of mean and variance, which improves it when dealing with over dispersed data (when variance is bigger than the mean). This is regularly the case with real data since they can be zero [27]. Therefore, negative binomial regression may be considered as a generalization of the Poisson regression.

For this model the value of *µ* differs from the one it takes on a Poisson regression, namely,

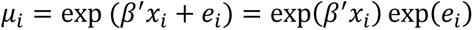

were *e*_*i*_ is a heterogeneity parameter and exp(*e*_*i*_) ∼Γ(*α*^−1^, *α*^−1^). The density function for *Y*_*i*_ is:

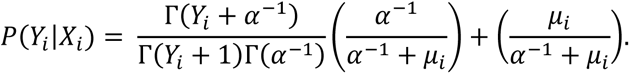

Finally, the Poisson-inverse Gaussian (PIG) regression is very similar to the negative binomial, but instead of combining the Poisson distribution with a Gamma distribution, it uses an inverse Gaussian distribution. This model is particularly good for data with a high initial peak and that might be skewed to the far right, o highly over dispersed data [28].

For the PIG regression *α* represents the dispersion parameter and can also be parametrized as *ϕ* = 1/*α*. Thereby, the probability distribution function is:

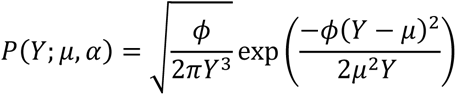

with *Y*, *µ*, *ϕ* > 0.

## 3. Results

### 3.1. Data imputation

The first obstacle to present itself was the presence of missing data on the climatic data set. This a fairly common difficulty when working with climatic data. A process of data imputation was necessary beforehand. For this purpose, the random forest regression method was used [29]. First, for every variable, a second meteorological station near Universidad del Valle without so many missing values was selected (Palmira or Jamundí), and then, the missing values of this new stations were replaced with the mean, and finally, a random forest regression model was trained using the secondary station to predict the missing values of the station at Universidad del Valle. As an example, in Fig 2 shows graphs for the original data and the completed version with imputation for mean temperature.

**Fig. 2.**
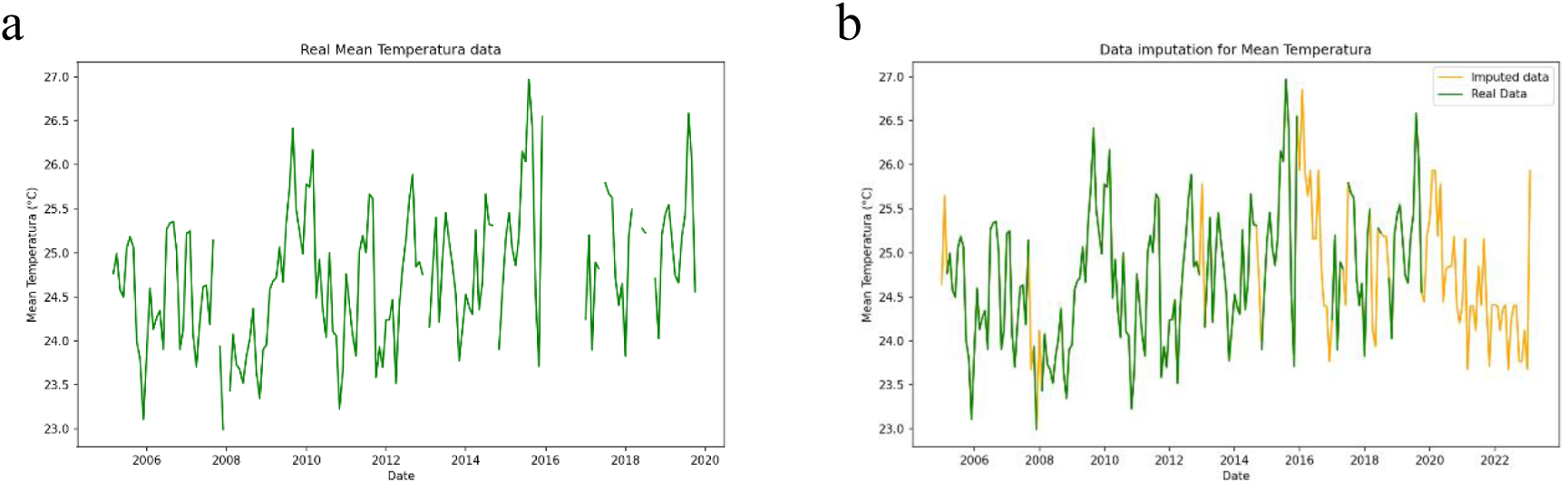
a) original data; b) data with imputation

It should be noted that information form 2005 up to 2022 was used in order to have more data for the regression model.

### 3.2. Lagged correlation analysis and election of variables

Fig 3 shows the time series of dengue incidence in Cali. It can be observed that dengue incidence is characterized by long periods of relative tranquility, where there are a few dengue cases, and some peaks where incidence shoots of and reaches much higher values. Bellow the relationship between this behavior and the aforementioned climatic variables.

**Fig. 3.**
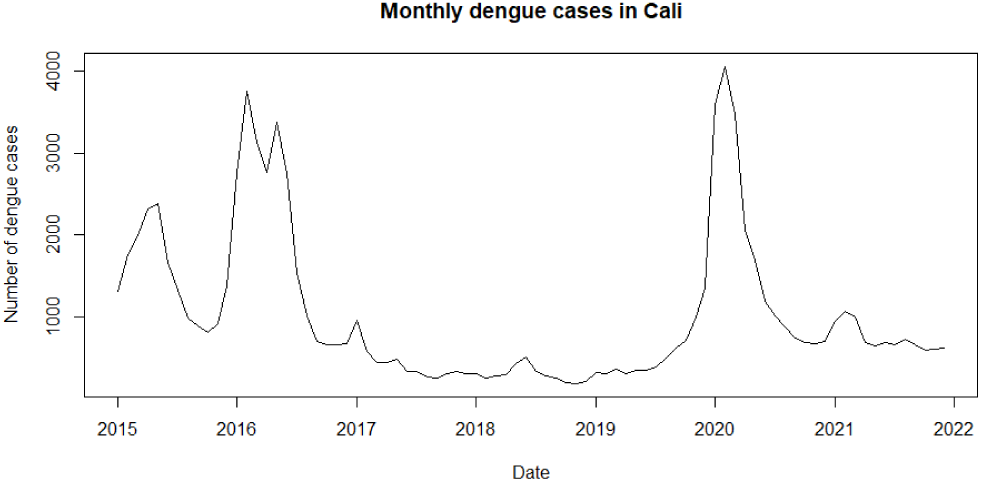
Monthly dengue cases in Cali

Using R, the correlation of each variable with dengue incidence was studied. However, the nature of the vector implies the need of considering a lag. In fact, later on we verify that the lagged variables show higher correlation with dengue incidence than without any lag. Fig 4 shows the lagged cross correlation analysis for all the climatic variables.

**Fig. 4.**
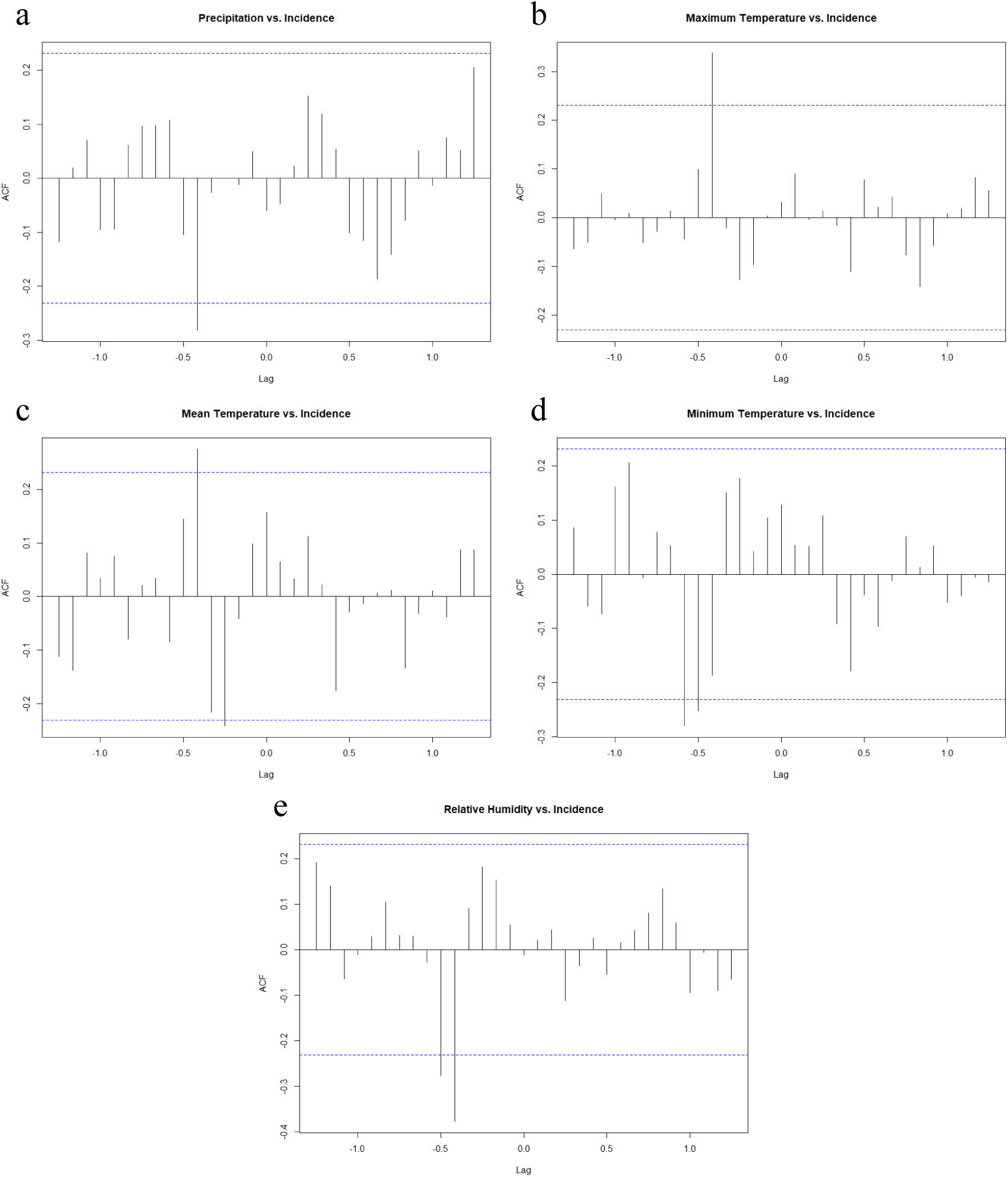
Lagged correlation analysis

For precipitation (Fig 4. a) there’s significant negative correlation at -0.4 lag, meaning between incidence and the precipitation of 5 months in the past. For maximum temperature (Fig. 4 b) the most significant correlation is around a -0.4 lag, resulting in a 4-month lag. The relation is direct. For average temperature (Fig. 4 c) there are two lags with significant correlation, a direct correlation with a 5-month lag and an inverse correlation with a 3-month lag. For minimum temperature the significant correlation is negative at a 7-month lag. And for relative humidity there is negative significant correlation at a 5-month lag. To summarize, the chosen variables after the lagged correlation analysis are Precipitation(t-5), Maximum Temperature(t-5), Mean Temperature(t-3), Mean Temperature(t-5), Minimum Temperature(t-7) and Relative Humidity(t-5). It must be noted that before doing this analysis the first difference of ever variables was calculated in order to guarantee stationarity and avoid spurious correlation.

### 3.3. Principal component analysis (ACP)

The multicollinearity problem must be addressed since, naturally, climatic variables are correlated amongst themselves. Table 1 shows de correlation matrix of the climatic variables.

**Table 1.** Correlation matrix of the climatic variables.

| Correlation matrix |  |  |  |  |  |  |  |
| --- | --- | --- | --- | --- | --- | --- | --- |
|  |  | Precipitation<br>(t-5) | Maximum<br>Temperature<br>(t-5) | Mean<br>Temperature<br>(t-5) | Mean<br>Temperature<br>(t-3) | Minimum<br>Temperature<br>(t-7) | Relative<br>Humidity<br>(t-5) |
| Correlation | Precipitation<br>(t-5) | 1,000 | -,388 | -,387 | ,012 | ,116 | ,555 |
|  | Maximum<br>Temperature<br>(t-5) | -,388 | 1,000 | ,766 | ,065 | -,054 | -,605 |
|  | Mean<br>Temperature<br>(t-5) | -,387 | ,766 | 1,000 | ,119 | ,096 | -,698 |
|  | Mean<br>Temperature<br>(t-3) | ,012 | ,065 | ,119 | 1,000 | ,238 | ,060 |
|  | Minimum<br>Temperature<br>(t-7) | ,116 | -,054 | ,096 | ,238 | 1,000 | ,174 |
|  | Relative<br>humidity (t-5) | ,555 | -,605 | -,698 | ,060 | ,174 | 1,000 |
| Sig.<br>(unilateral) | Precipitation<br>(t-5) |  | <.001 | <.001 | ,460 | ,167 | <.001 |
|  | Maximum<br>Temperature<br>(t-5) | ,000 |  | ,000 | ,295 | ,327 | ,000 |
|  | Mean<br>Temperature<br>(t-5) | ,000 | ,000 |  | ,161 | ,212 | ,000 |
|  | Mean<br>Temperature<br>(t-3) | ,460 | ,295 | ,161 |  | ,023 | ,309 |
|  | Minimum<br>Temperature<br>(t-7) | ,167 | ,327 | ,212 | ,023 |  | ,073 |
|  | Relative<br>humidity (t-5) | ,000 | ,000 | ,000 | ,309 | ,073 |  |

Fig 5 shows the associated variance to each of the principal components. According to this, we pick the first to components which explain 67,3% of the variance.

**Fig. 5.**
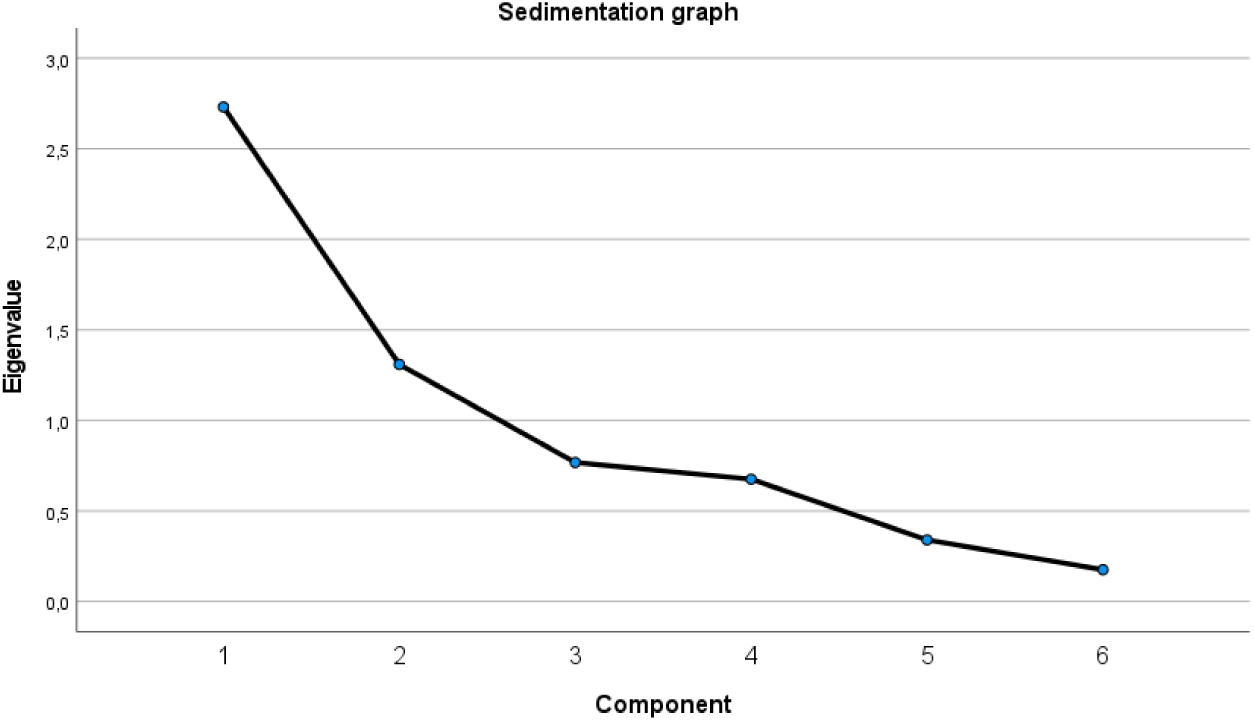
Sedimentation graph of the ACP

Fig 6 reveals that the variables can be separated in two groups. Therefore, instead of using the principal components as predictor variables, we choose one variable for each group making the regression easier to understand and conclude. The selected variables are Mean Temperature(t-5) and Mean Temperature(t-3).

**Fig. 6.**
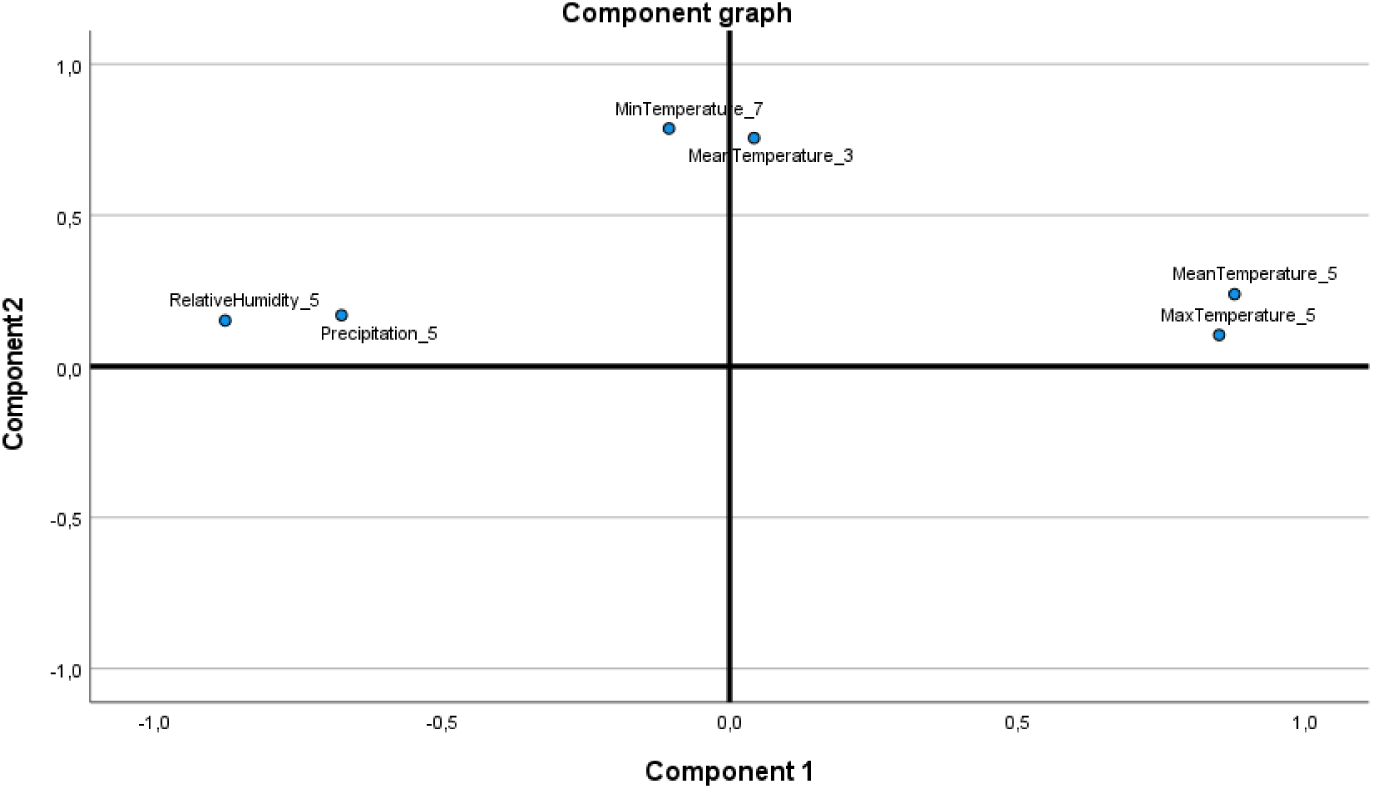
Principal components in rotated space

### 3.4. Regression models

Table 2 presents a comparison of the three regression models used in this work including deviance, Akaike information criterion (AIC) and the p-values of the adjusted models for every variable. For every model the best variable choice was both mean temperature lag variables and their interaction.

**Table 2.** Comparison of the regression models.

| Model | Residual deviance | AIC | P-VALUES |  |  |  |
| --- | --- | --- | --- | --- | --- | --- |
|  |  |  | Intercept | Mean Temperature (t-5) | Mean temperature (t-3) | Interaction |
| Poisson | 34224.0 | 34826.0 | 2.00E-16 | 2.00E-16 | 2.00E-16 | 2.00E-16 |
| Negative binomial | 76.388 | 1095.80 | 0.00898 | 0.00893 | 0.01 | 0.01249 |
| PIG | 1075.814 | 1085.81 | 0.0439 | 0.0423 | 0.0463 | 0.0539 |

Clearly, the Poisson model is not good, probably because dengue incidences is over dispersed, its mean is 981.6 and its variance is 950046.6. Therefore, this model was discarded. Regarding the negative binomial and the PIG models, the negative binomial is chosen because its deviance is better and shows a slightly better adjustment to the data as seen in the p-values. As Table 3 shows, all the variables and the intercept were significant for the negative binomial model.

**Table 3.**
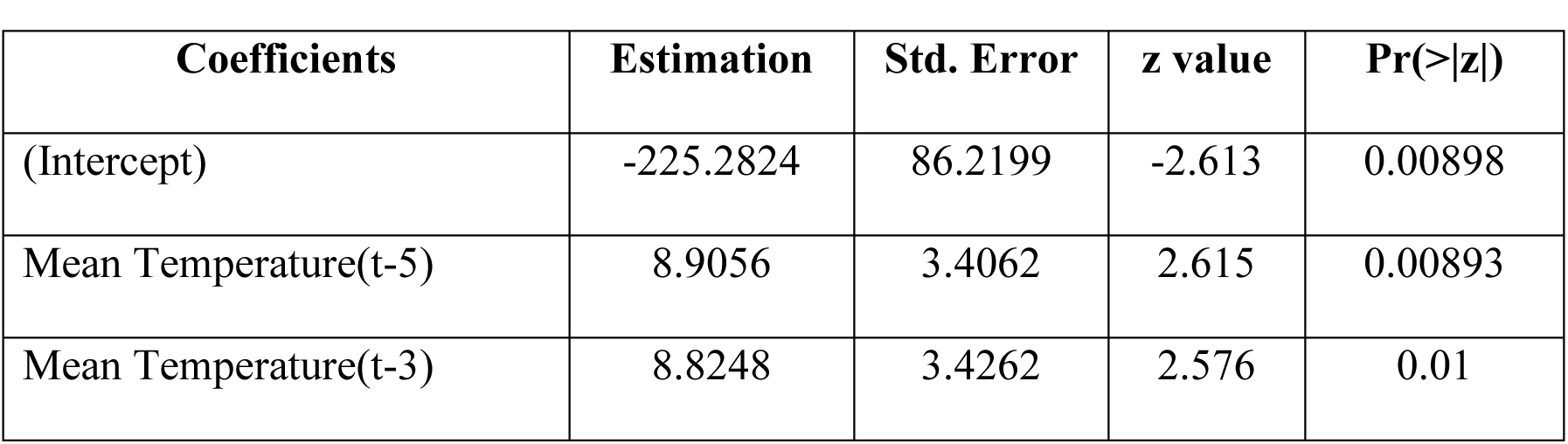
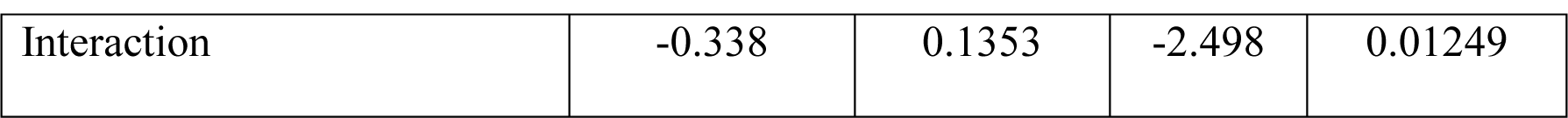
Coefficients of the negative binomial model.

We can also verify that the data resemble a negative binomial distribution as Fig 7 shows.

**Fig. 7.**
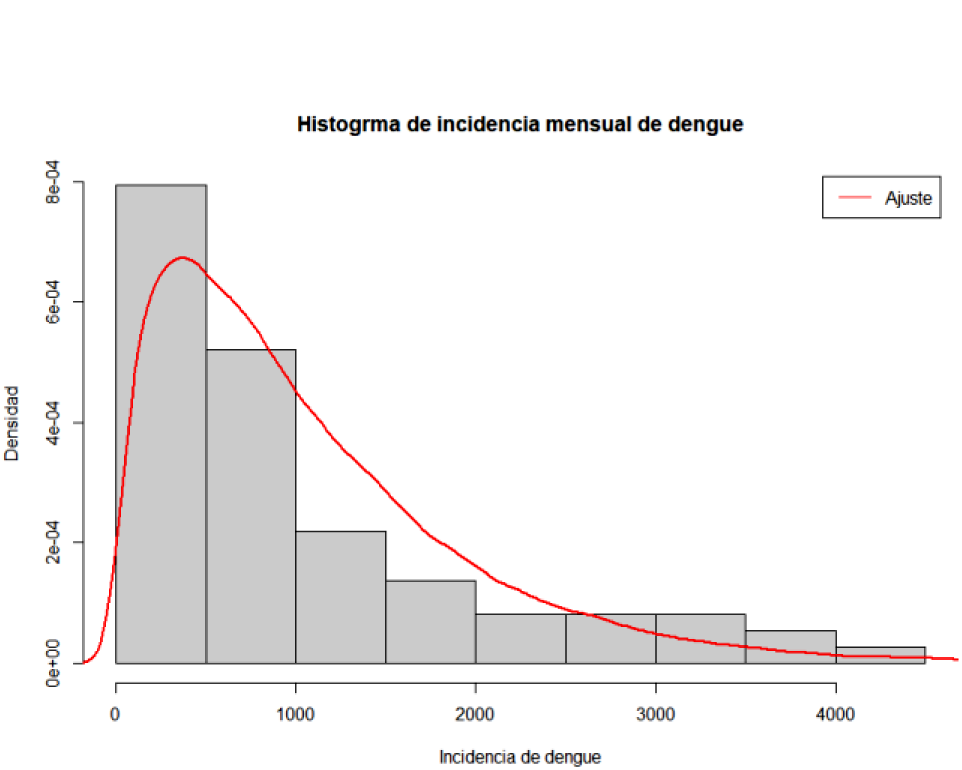
Frequency distribution

The resulting model can be expressed as follows:

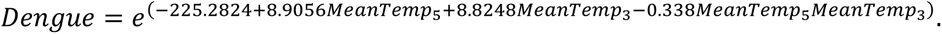

Fig 8 displays both real data and data predicted by the negative binomial regression model. The fit is good since the predicted values (red) resemble the real data (black) in the time series as well as the histogram (Fig 9). However, it’s clear that the fit could be better, for example, the peaks of 2016 and 202 aren’t predicted correctly, while the dengue cases on the low incidence seasons are overestimated. It must be kept in mind that 2020 was an atypical year due to the Covid-19 pandemic.

**Fig. 8.**
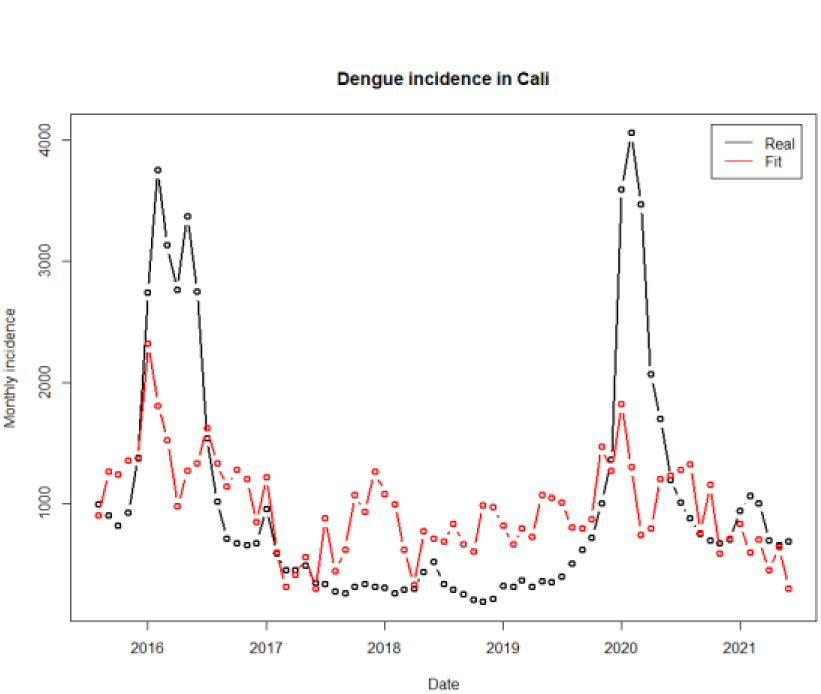
Adjusted regression model for dengue incidence

**Fig. 9.**
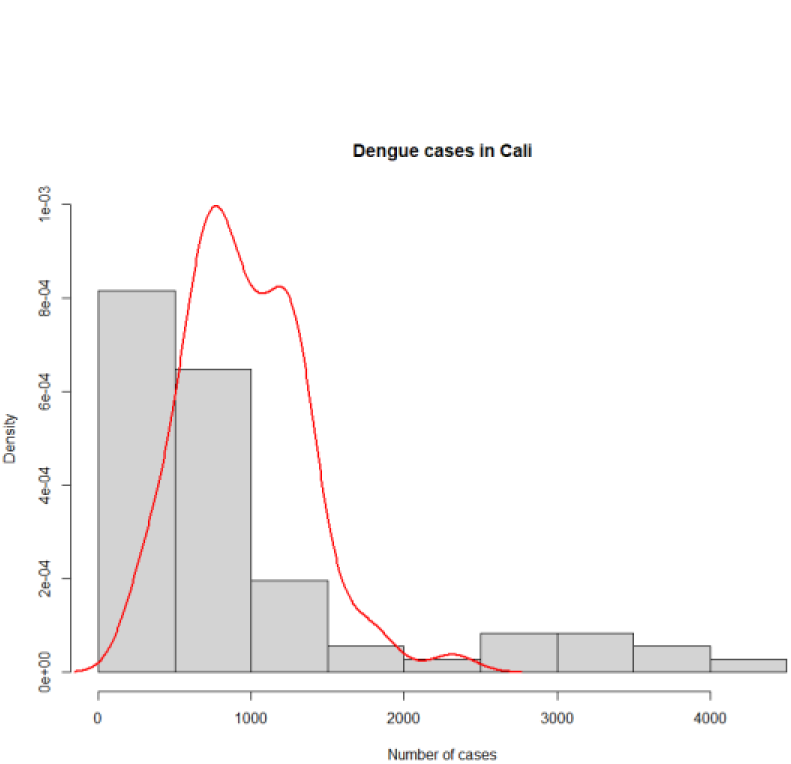
Histogram of dengue incidence

Knowing that the average temperature for the data set is 25°C, the coefficients can be used for the following interpretations: If the average monthly temperature in Cali increases by one degree, a 45% increase in dengue cases is expected after 3 months. Additionally, after 5 months, the same increase of one degree will imply a 64% increase in dengue incidence. This is particularly worrying considering the temperature rise tendencies because of global warming and the recent arrival of El Niño phenomenon.

## 4. Discussion

The results obtained on the data imputation process shows that this methodology is a feasible and efficient way of estimating missing data.

The negative binomial regression model was found to be the best for predicting monthly dengue incidence in Cali using mean temperature with 3 and 5 months lags. This implies that the effect of a change in temperature takes about 3 months to be visible in dengue incidence. This could be attributed to the biological processes of *Aedes aegypti* such as maturation and reproduction which are known to be affected by the climatic variables. Taking into consideration that the life span of a female mosquito is about 6 weeks, we can conclude that it takes at least two generation of mosquitoes for the dengue incidence to be affected by a change in temperature.

In other matters, the absence of other climatic variables from the regression model does not necessarily imply that they have no effect over dengue incidence, seeing that the naturally have a big correlation with temperature. For instance, both precipitation and relative humidity are inversely proportional to temperature, as the ACP shows.

Forecasts are worrisome, because an increase in average temperature could transform into a significant increase in dengue cases having serious repercussions on public health.

Finally, its recommended to deepen on this type of predictive studies. This research has some limitations that can be improved upon on further work. Firstly, there isn’t a lot of climatic data. Only one meteorological station with enough data was found, and even then, this data was incomplete and hence an imputation processes was necessary. Secondly, expanding the time period of the study is recommended in order to obtain more robust results, but this will imply recollecting more data for both dengue and the climatic variables. And finally, this work didn’t take into consideration the autoregressive component of dengue (present dengue incidence depends on the incidence of past months). This could lead to more accurate predictions.

## Data Availability

All the data used for this article is available at the Dryad repository following thins link: https://doi.org/10.5061/dryad.0zpc8675h

